# Crescentic Glomerulonephritis: What’s different in South Asia? - A Single Center Observational Cohort Study

**DOI:** 10.1101/2020.06.12.20129122

**Authors:** Suceena Alexander, Sabina Yusuf, Gautham Rajan, Elenjickal Elias John, Sanjeet Roy, VC Annamalai, Athul Thomas, Jeethu Joseph Eapen, Anna T. Valson, Vinoi George David, Santosh Varughese

## Abstract

**Background:** The spectrum and outcomes of crescentic glomerulonephritis in South Asia is vastly different from that reported worldwide and there is a paucity of information.

**Methods:** It was an observational cohort study of renal biopsies done in the largest tertiary center in South India over a period of 10 years with ≥50% crescents on histology.

**Results:** A total of 8645 kidney biopsies were done at our center from January 2006 to December 2015, and 200 (2.31%) were crescentic glomerulonephritis. Patients were categorized into three etiological groups - anti-GBM (type I), immune complex (type II) and pauci-immune (type III). The most common was type II (96, 46.5%), followed by type III (73, 38%) and then type I (31, 15.5%). Female preponderance was seen across all three types. About half of all the three types presented with recent onset hypertension. Type II had the highest median proteinuria (4.2 (2.1-6) g/day, p=0.06) and the median eGFR was lowest in type I (5 (4-8) ml/min/1.73m^2^, p<0.001). Among type III, ANCA associated vasculitis (AAV) was seen only in about half of the patients. Nearly one third patients with type I was also positive for ANCA making them ‘double positive’. Severe glomerular insults like tuft necrosis and chronicity as evidenced by moderate to severe interstitial fibrosis was a predominant feature of type I.

**Conclusions:** ANCA negative vasculitis as well as double positive types are reported for the first time from South-Asia. Prevalence of ANCA negative vasculitis (type III subgroup) was much higher in our population. Renal survival was significantly worse in type I & III compared to type II. Types I/III, moderate to severe IFTA, presence of oliguria/anuria and increasing percentage of crescents in renal biopsy were significant predictors of dialysis dependence at index visit or of end stage kidney disease at follow-up in our cohort.

## INTRODUCTION

Crescentic glomerulonephritis (Cr.GN) is characterized by the presence of extensive glomerular crescents (usually greater than 50%) as the principal histologic finding. Because it often clinically presents with a rapid decline in kidney function, it is also known as rapidly progressive glomerulonephritis (RPGN). It can complicate any glomerular disease^1^ and is present in approximately 4% to 10% of native kidney biopsies.^2^ The natural course of the disease renders it a “medical emergency”, as the decline in renal function is relentless and leads to end stage renal failure in a few weeks or months.^3^ Understanding the clinical presentation, natural history and outcomes of crescentic glomerulonephritis is of major concern for nephrologists worldwide. Various studies have been conducted worldwide and epidemiologic data are available from large national kidney biopsy registries including those from the United States^4^, China^2^, Japan^5^, Spain^6^ and Saudi Arabia.^7^ An important thing to realise here is that pathogenesis of glomerular diseases involves a complex and as yet incompletely understood interplay between epigenetic, immunoregulatory, hormonal, and environmental factors on a background of genetic predisposition.^8^ This translates into a broad spectrum of disease presentation, a variable tempo of progression and heterogenous outcomes which are evident from these studies. To this end, exploring the features of crescentic glomerulonephritis in our population, which is yet another genetically and demographically diverse group of individuals may help to provide useful insights into the causality, and predictors of severe outcomes.

## METHODS

This was an observational retrospective cohort study done in outpatient and inpatient services of the Department of Nephrology in our hospital. We included all patients (≥18 years) who underwent native renal biopsy at our centre between January 2006 to December 2015 and had ≥50% crescents on histology. Data on their demographic profile, clinical features, biochemical parameters, histopathology, treatments, morbidity and mortality were retrieved from the electronic patient records (Clinical WorkStation) maintained in the hospital. Follow-up clinical and outcome data with regards to their serum creatinine, dialysis requirement, and complications were collected for each review visit till August 2016. During the index and follow up visits, patients were classified into chronic kidney disease (CKD) stages as per estimated glomerular filtration rate (eGFR) calculated by CKD-EPI equation.^9^

Cr.GN was defined as the presence of ≥50% glomerular crescents as the principal histologic finding. Patients were categorized into three groups on the basis of etiology of crescentic GN-anti-GBM crescentic GN (type I Cr.GN), immune complex crescentic GN (type II Cr.GN), pauci-immune crescentic GN (type III Cr.GN). Microhematuria was defined as >5 red blood cells (RBC’s) per high power field. Proteinuria was assessed from 24-hour timed collection as is the standard practice in our center. Interstitial fibrosis and tubular atrophy (IFTA) was classified into: focal-<25%, moderate-25% to 50% and severe->50%. Qualitative and semiquantitative determination of anti-nuclear antibody (ANA) in serum was done manually by EUROIMMUN Mosaic Hep-20-10 indirect immunofluorescence test (IIFT). The test was done with a sample dilution starting point of 1:100. It is graded on a scale of 1+ to 5+. The sensitivity of the test was 100% with a specificity of 96%. Quantitative determination of anti-double stranded DNA (anti-dsDNA) in serum was done by Anti-dsDNA-NcX ELISA (IgG). The upper limit of the normal range (cut-off) was 100 IU/ml. Anti-neutrophil cytoplasmic antibodies (ANCA) were determined by measuring anti-myeloperoxidase (anti-MPO) and anti-proteinase3 (anti-PR3). Quantitative determination of anti-MPO was done by Anti-Myeloperoxidase ELISA (IgG) test kit. The upper limit of the normal range (cut-off) is 20 RU/ml. The ELISA had a sensitivity of 93.3% and a specificity of 99.8% with regard to IIFT. Quantitative determination of anti-PR3 was done by Anti-PR3-hn-hr ELISA (IgG). The upper limit of the normal range (cut-off) was 20 RU/ml. The ELISA had a sensitivity of 94% and a specificity of 99%. The tests kits for antibodies were from EUROIMMUN, Luebeck, Germany. Quantitative determination of complement factors (C3 and C4) was done by means of endpoint nephelometry on the BN ProSpec System by Siemens Health Care Diagnostics Products, Marburg, Germany. Antisera used were liquid animal sera produced by immunization of rabbits with highly purified human complement factors (C3c or C4). The following reference intervals applied for serum samples from healthy adults: C3/C3c: 0.9-1.8 g/L, C4/C4c: 0.1-0.4 g/L.

### Statistical Analysis

Data are presented as mean ± standard deviation or medians (interquartile range) or frequency & percent according to the types and distribution of variables. Differences among groups of normally distributed variables were analyzed by t test or one-way analysis of variance (ANOVA). Post-hoc comparisons were performed using t Test with Bonferroni correction. Differences among groups of non-parametric variables were analyzed by Mann– Whitney U-test or the Kruskal-Wallis Test. Categorical variables were compared using chi-squared or Fisher’s exact test. Multivariable logistic regression was used to identify predictors of end stage kidney disease (ESKD). Statistical calculations were performed using SPSS software for Windows, version 21.0 (SPSS Inc., Chicago, IL) and graphs were made using Graph Pad Prism 7.0e (Graph Pad Software Inc., San Diego, CA). A *P* value of <0.05 was taken as significant.

## RESULTS

### Demography

A total of 8645 kidney biopsies were done at our center from January 2006 to December 2015, out of which 200 were found to have Cr.GN (2.31%). The most common cause of Cr.GN was type II (96, 46.5%), followed by type III (73, 38%) and least common was type I (31, 15.5%). The various etiologies of Cr.GN are depicted in Figure 1. Females constituted 60% of the patients with a female: male ratio of 1.5:1. Female preponderance was seen across all three types of Cr.GN. The mean age of presentation was 40.6±14.6 years with the highest mean age seen in type III Cr.GN. Demographic and baseline clinical and laboratory parameters of the study cohort are summarized in table 1.

**Table 1.** Demography, baseline clinical and laboratory characteristics

| Characteristic | Entire Cohort<br>(n=200) | Type I Cr. GN<br>(Anti-GBM)<br>(n=31) | Type II Cr. GN<br>(Immune Complex)<br>(n=93) | Type III Cr. GN<br>(Pauci-Immune)<br>(n=76) | P value |
| --- | --- | --- | --- | --- | --- |
| Age (years, mean±SD) | 40.6±14.6 | 37.5±12.2 | 37.8±14.6 | 45.3± 14.3 | **0.03<sup>@</sup>, 0.003<sup>#</sup>** |
| Gender (male:female (ratio)) | 80:120 (0.67) | 12:19 (0.63) | 36:57 (0.63) | 32:44 (0.73) | 0.89 |
| Renal symptoms |  |  |  |  |  |
| Oliguria (n, %) | 112 (56) | 23 (74.2) | 50 (53.8) | 39 (51.3) | 0.08 |
| Anuria (n, %) | 20 (10) | 5 (16.1) | 4 (4.3) | 11 (4.5) | **0.03** |
| Visible hematuria (n, %) | 25 (12.5) | 6 (19.4) | 9 (9.7) | 10 (13.2) | 0.38 |
| Non-visible hematuria (n, %) | 190 (95) | 29 (93.5) | 90 (96.8) | 71 (93.4) | 0.55 |
| Hypertension (n, %) | 100 (50) | 15 (48.4) | 51 (54.8) | 34 (44.7) | 0.42 |
| Uremic symptoms (n, %) | 84 (42) | 20 (64.5) | 30 (32.3) | 34 (44.7) | **0.006** |
| Extra-renal symptoms |  |  |  |  |  |
| Skin lesions (n, %) | 24 (12) | 0 | 16 (17.2) | 8 (10.5) | **0.006** |
| Arthritis (n, %) | 40 (20) | 6 (19.4) | 18 (19.4) | 16 (21.1) | 0.96 |
| Hemoptysis (n, %) | 19 (9.5) | 4 (12.9) | 0 | 15 (19.7) | **<0.001** |
| Diabetes Mellitus (n, %) | 14 (7) | 0 | 5 (5.4) | 9 (11.8) | **0.03** |
| Hemoglobin (g/dL, mean±SD) | 8.6±2.1 | 7.6±1.9 | 9±2 | 8.5±2.1 | **0.003<sup>S</sup>** |
| Total leucocyte (cells*10<sup>9</sup>/L, mean±SD) | 9.9±4.5 | 9.3±3.9 | 9.8±5.2 | 10.3±3.7 | 0.57 |
| Serum cholesterol (mg/dL, mean±SD, n) | 199.7±73.8<br>(123) | 205.3±101.8<br>(15) | 213.9±73.2 (62) | 178.8±59.6 (46) | **0.04<sup>#</sup>** |
| Serum Albumin (mg/dL, mean±SD) | 3±0.7 | 3.2±0.6 | 2.8±0.8 | 3.1±0.5 | **0.008<sup>S</sup>, 0.005<sup>#</sup>** |
| 24-hour proteinuria (g/day, median (IQR), n) | 3.5 (1.8-5.6)<br>(187) | 3.3 (2-5.8) (26) | 4.2 (2.1-6) (89) | 2.8 (1.3-4.3) (72) | **0.06** |
| Serum creatinine (mg/dL, mean±SD) | 6.9±4.4 | 10.6± 5.5 | 5.1±3.2 | 7.6±4.1 | **<0.001<sup>@S</sup>, 0.001<sup>#</sup>** |
| CKD-EPI eGFR<br>(ml/min/1.73m<sup>2</sup>, median (IQR)) | 9 (5-16.8) | 5 (4-8) | 14 (8-25.5) | 7 (5-12) | **<0.001<sup>@#</sup>** |
| CKD stages at baseline |  |  |  |  |  |
| Stage 2 Yes (n/N, %) | 7 (3.5) | 0 | 6 (6.5) | 1 (1.3) | **<0.001** |
| Stage 3 Yes (n/N, %) | 19 (9.5) | 1 (3.2) | 13 (14) | 5 (6.6) |  |
| Stage 4 Yes (n/N, %) | 34 (17) | 0 | 24 (25.8) | 10 (13.2) |  |
| Stage 5 Yes (n/N, %) | 52 (26) | 6 (19.4) | 21 (22.6) | 25 (32.9) |  |
| Stage 5D Yes (n/N, %) | 88 (44) | 24 (77.4) | 29 (31.2) | 35 (46.1) |  |
| Serum complements (mg/dL) |  |  |  |  |  |
| Low C3 (n/N, %) | 84/196 (42.9) | 7/31 (22.6) | 59/90 (65.6) | 18/75 (24) | **<0.001** |
| Low C4 (n/N, %) | 21/196 (10.7) | 0 | 20/90 (22.2) | 1/75 (1.3) | **<0.001** |
| Serology |  |  |  |  |  |
| ANA Yes (n/N, %) | 51/176 (29) | 7/31 (22.6) | 38/80 (47.5) | 6/65 (9.2) | **<0.001** |
| Anti- dsDNA Yes (n/N, %) | 25/146 (17.1) | 0 | 25/75 (33.3) | 0 | **<0.001** |
| ANCA Yes (n/N, %) | 54 (31.8) | 10/31 (32.3) | 6/64 (9.4) | 38/76 (50) | **<0.001** |
| Anti-MPO-ANCA Yes (n/N, %) | 28/143 (19.6) | 7/25 (28) | 3/50 (6) | 18/68 (26.5) | **<0.001** |
| Anti-PR3-ANCA Yes (n/N, %) | 26/143 (18.2) | 3/25 (12) | 3/50 (6) | 20/68 (29.4) | **<0.001** |

Abbreviations: eGFR, estimated glomerular filtration rate (calculated using the CKD-EPI, Chronic Kidney Disease Epidemiology
Collaboration (CKD-EPI) formula); C3, complement C3; C4, complement C4; ANA, anti-nuclear antibody; Anti- dsDNA, anti-double stranded DNA antibody, ANCA, anti-neutrophil cytoplasmic antibody; MPO; myeloperoxidase; PR3, proteinase 3
P value is significant at <0.05 between <sup>@</sup> Type I and Type III, <sup>#</sup> Type II and Type III, <sup>S</sup>Type I and Type II analyzed by One-way ANOVA with Bonferroni correction.

**Figure 1.**
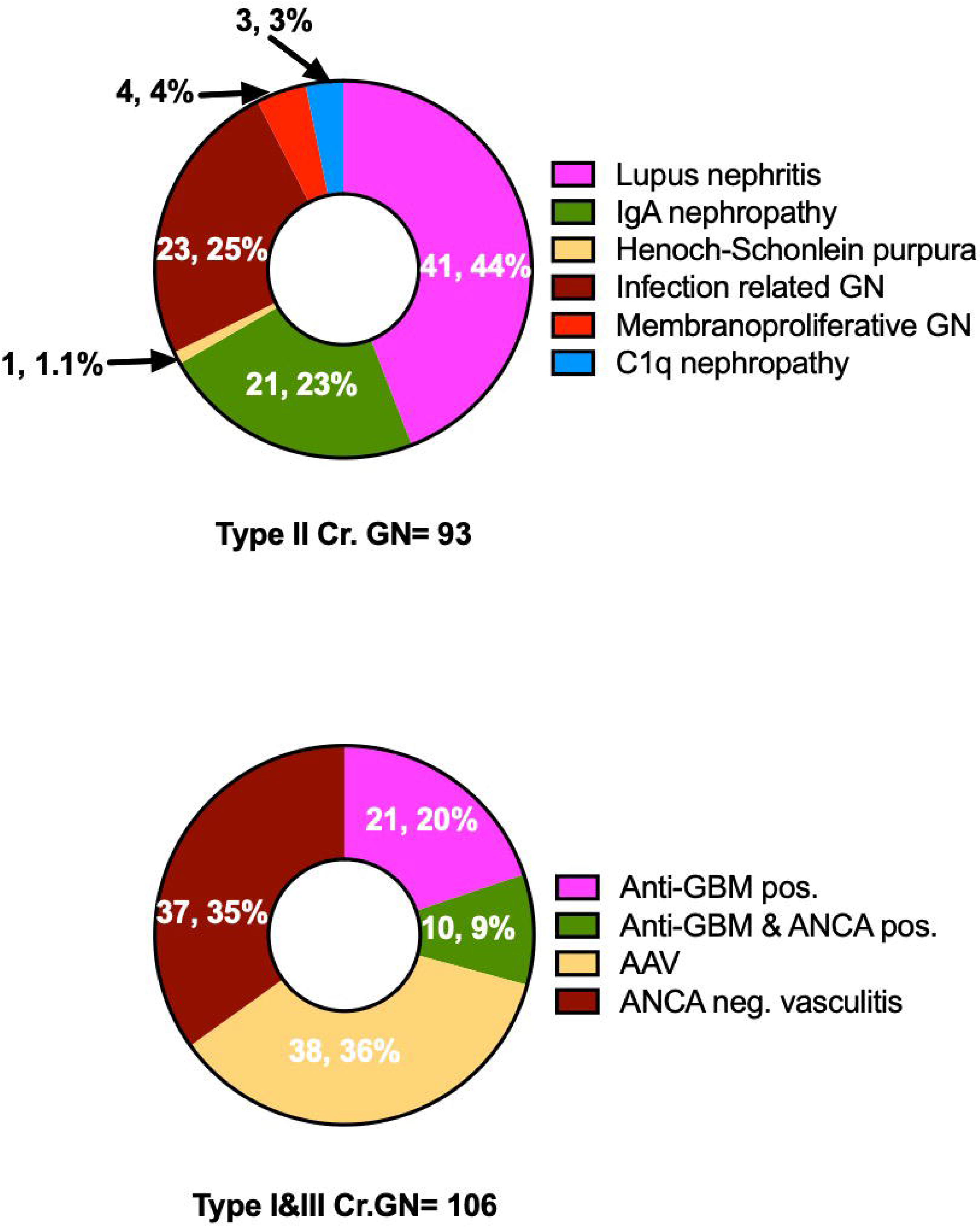
Etiologies of crescentic glomerulonephritis

### Clinical and laboratory features

Non-visible hematuria was near universal (95%). More than half of the patients (56%) were oliguric at presentation. Anuria at presentation was seen in 10% patients. Oliguria and anuria were more common in type I Cr.GN patients (oliguria in 74%; p=0.08 and anuria in 16%; p=0.04) who also had significantly more uremic symptoms (64%, p=0.006). About half of all the three types presented with recent onset hypertension. Among extra-renal symptoms, skin lesions and arthritis were rare in type I but hemoptysis was seen only in type I & III Cr.GN. Type II Cr.GN had the highest median proteinuria (4.2 (2.1-6) g/day, p=0.06), lowest serum albumin (2.8±0.8 g/dL, p <0.001) and highest serum cholesterol levels (214±73 mg/dL, p=0.04). The median eGFR was 9 (5-17) ml/min/1.73m^2^ and was lowest in type I Cr.GN (5 (4-8) ml/min/1.73m^2^, p<0.001) with the lowest mean Hb (7.6±2 g/dL, p=0.003). Serum complement levels were low in nearly 50% patients with Cr.GN (low C3 in 43% and low C4 in 11%), predominantly in those with type II Cr.GN (p <0.001). Interestingly, low C3 levels were also found in about 25% patients with either type I or type III Cr.GN. Low C4 levels, on the other hand was seen only in type II Cr.GN. Though ANA was positive to various extents in all types of Cr.GN (23% in type I, 9% in type II & 47% in type III), dsDNA positivity was seen only type II Cr.GN (33%). Among type III Cr.GN, ANCA associated vasculitis (AAV) was seen only in 51% patients (anti-MPO 26%; anti-PR3 29%). Nearly one third patients with type I Cr.GN (32%) showing linear staining in immunofluorescence (IF) were also positive for ANCAs (anti-MPO 28%; anti-PR3 29%) making them ‘double positive Cr.GN’.

### Histopathological features

Characteristic histopathological features noted in kidney biopsies have been summarized in table 2. The highest percentage of glomeruli with crescents was seen in type I (83±17%, p=0.001) followed by type III (74±19%) and type II Cr.GN (69±18%). The most common type of crescents in all three groups was fibrocellular. Glomerular proliferative lesions as evidenced by mesangial hypercellularity (68%, p=0.02), endocapillary proliferation (100%) and neutrophilic exudation (50%, p=0.01), were predominantly seen in type II Cr.GN. Severe glomerular insults like tuft necrosis was a predominant feature of type I (32%, p=0.002) and type III Cr.GN (25%). Chronicity as evidenced by moderate to severe IFTA was most severe in type I Cr.GN (68%; p=0.02).

**Table 2.** Histopathology characteristics

| Characteristic | Entire Cohort<br>(n=200) | Type I Cr. GN<br>(Anti-GBM)<br>(n=31) | Type II Cr. GN<br>(Immune Complex)<br>(n=93) | Type III Cr. GN<br>(Pauci-Immune)<br>(n=76) | P value |
| --- | --- | --- | --- | --- | --- |
| Number of glomeruli (median (IQR)) | 9 (6-12) | 11 (6-15) | 9 (6-11) | 9.5 (6-12) | 0.46 |
| Number of sclerosed glomeruli (median (IQR)) | 1 (0-3) | 0 (0-6) | 1 (0-3) | 1 (0-4) | 0.23 |
| Crescents (%) | 73.2±18.9 | 83±16.7 | 69.2±18.1 | 74.2±19.4 | **0.001<sup>s</sup>** |
| Predominant type |  |  |  |  | ] 0.17 |
| Cellular (n, %) | 49 (24.5) | 5 (16.1) | 22 (23.7) | 22 (29) |  |
| Cellular to fibro-cellular (n, %) | 40 (20) | 10 (32.3) | 18 (19.3) | 12 (15.8) |  |
| Fibrocellular (n, %) | 88 (44) | 10 (32.3) | 46 (49.5) | 32 (42.1) |  |
| Fibrous (n, %) | 23 (11.5) | 6 (19.4) | 7 (7.5) | 10 (13.2) |  |
| Glomerular lesions |  |  |  |  |  |
| Mesangial proliferation Yes (n, %) | 116 (58) | 13 (41.9) | 63 (67.7) | 40 (52.6) | **0.02** |
| Inter-capillary mesangial sclerosis Yes (n, %) | 16 (8) | 6 (19.3) | 3 (3.2) | 7 (9.2) | **0.02** |
| Endocapillary proliferation Yes (n, %) | 79 (100) | - | 79 (100) | - |  |
| Neutrophilic infiltration Yes (n, %) | 79 (39.5) | 9 (29) | 47 (50.5) | 23 (30.3) | **0.01** |
| Tuft necrosis Yes (n, %) | 37 (18.5) | 10 (32.2) | 8 (8.6) | 19 (25) | **0.002** |
| Glomerular thrombosis Yes (n, %) | 3 (1.5) | 0 | 3 (3.2) | 0 | **0.1** |
| Interstitial |  |  |  |  |  |
| IFTA Moderate/Severe Yes (n, %) | 100 (50) | 21 (67.7) | 38 (40.9) | 41 (53.9) | **0.02** |
| Vascular |  |  |  |  |  |
| Necrosis Yes (n, %) | 7 (3.5) | 1 (3.2) | 3 (3.2) | 3 (3.9) | 0.96 |
| Arterio(lo)sclerosis Yes (n, %) | 55 (23) | 12 (38.7) | 27 (29) | 16 (21.1) | 0.95 |

Abbreviations: IFTA, interstitial fibrosis and tubular atrophy.
P value is significant at <0.05 between @ Type I and Type III, # Type II and Type III, <sup>s</sup>Type I and Type II analyzed by One-way ANOVA with Bonferroni correction.

### Treatment differences

Standard treatment protocols as per KDIGO 2012^10^ guidelines were followed for all patients. Nearly half of the patients required dialysis at presentation, with significantly more in type I and type III Cr.GN (81% & 62% respectively, p<0.001). About 1/5^th^ of patients received therapeutic plasma exchange (PLEX) and this was significantly more in type I and type III Cr.GN (42% & 26% respectively, p<0.001). The frequency and indications of PLEX are summarized in table 3. Immunosuppression with/without PLEX was given in 90% of patients with significantly more in type II & III Cr.GN (92% & 91% respectively, p<0.001).

**Table 3.** Treatment characteristics

| Characteristic | Entire Cohort<br>(n=200) | Type I Cr. GN<br>(Anti-GBM)<br>(n=31) | Type II Cr. GN<br>(Immune Complex)<br>(n=93) | Type III Cr. GN<br>(Pauci-Immune)<br>(n=76) | P value |
| --- | --- | --- | --- | --- | --- |
| Immunosuppression (IS) Yes (n, %) | 177 (88.5) | 22 (71) | 86 (92.5) | 69 (90.8) | **0.01** |
| Steroids alone Yes (n, %) | 43 (21.5) | 2 (35.5) | 29 (31.2) | 12 (15.8) | **0.003** |
| Steroids plus cyclophosphamide Yes (n, %) | 100 (50) | 18 (58.1) | 33 (35.4) | 49 (64.5) | **0.001** |
| Steroids plus other IS Yes (n, %) | 33 (16.5) | 1 (3.2) | 24 (25.8) | 8 (10.5) |  |
| PLEX with IS Yes (n, %) | 37 (18.5) | 12 (38.7) | 5 (5.4) | 20 (26.3) | **<0.001** |
| without IS Yes (n, %) | 1 (0.5) | 1 (3.2) | 0 | 0 |  |
| PLEX indications |  |  |  |  |  |
| Hemoptysis Yes (n, %) | 2 (1) | 1 (3.2) | 0 | 1 (1.3) | **<0.001** |
| Renal failure Yes (n, %) | 26 (13) | 10 (32) | 5 (5.4) | 11 (14.5) |  |
| Both Yes (n, %) | 11 (5.5) | 3 (9.7) | 0 | 8 (10.5) |  |
| Hemodialysis (n, %) | 104 (52) | 25 (80.6) | 32 (34.4) | 47 (61.8) | **< 0.001** |

Abbreviations: PLEX, plasma exchange; IS, immunosuppression.
P value is significant at <0.05 between <sup>@</sup> Type I and Type III, <sup>#</sup> Type II and Type III, <sup>s</sup> Type I and Type II analyzed by One-way ANOVA with Bonferroni correction.

### Patient and renal outcomes

The mean follow-up period was 9.4±15 months. Table 4 summarizes the outcomes observed in three types of Cr.GN. In the entire cohort, nearly half (45%) developed ESKD requiring renal replacement therapy. This was significantly more in type I (77%, p<0.001). Overall mortality rate was 4% and did not vary significantly between the groups. Sepsis was found to be the most common cause of death.

**Table 4.**
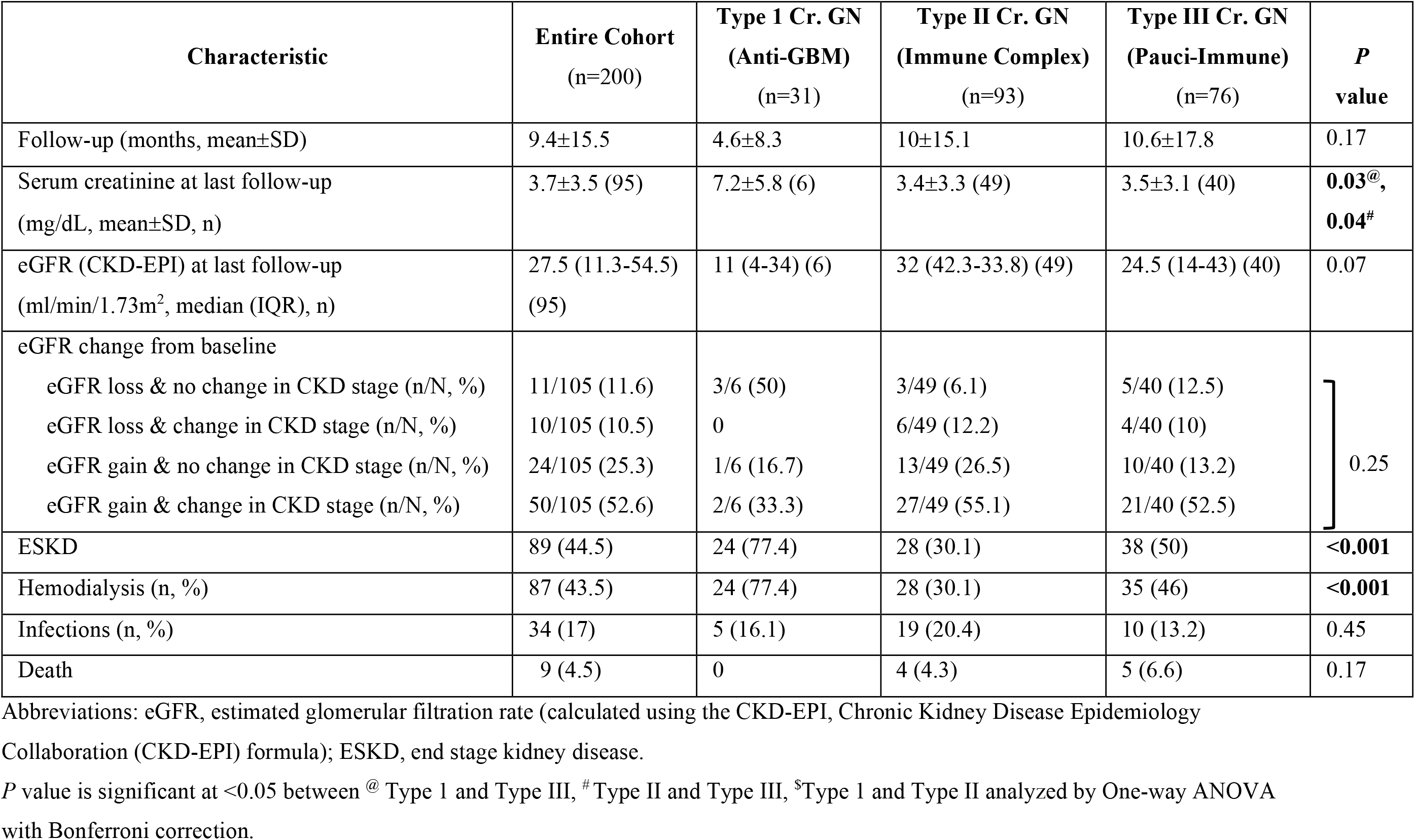
Outcomes at follow-up

### Type III Cr.GN: AAV vs. ANCA negative vasculitis

The group of patients with type III Cr.GN was subdivided into AAV (38, 51%) and ANCA negative (37, 49%) vasculitis. Characteristic features of the two groups are summarized in Table 5. AAV was associated with a significantly higher mean age (p=0.01), presence of extra renal manifestations of fever (50%, p=0.02), hemoptysis (39%, p<0.001) leukocytosis (11.2±4.5 cells* 10^9^/L, p=0.03), severe histology as evidenced by fibrous/ fibrocellular crescents (84%, p=0.009) & tuft necrosis (37%, p=0.02) and greater requirement of PLEX (45%, p<0.001). However, the median 24-hour proteinuria (3.7 (2.1-6.7) g/day, p=0.001) and serum cholesterol (206.7±61 mg/dL, p=0.003) was higher in ANCA negative vasculitis. Rates of ESKD and renal survival did not differ between the two groups (Table 5 & Figure 2).

**Table 5.** Type III Cr.GN: Characteristics of AAV and ANCA negative vasculitis

| Characteristic | ANCA neg. vasculitis(n=37) | AAV (n=38) | P value |
| --- | --- | --- | --- |
| Age (years, mean±SD) | 41±13.3 | 49.4±14.4 | **0.01** |
| Gender (male:female (ratio)) | 15:22 (0.68) | 16:22 (0.73) | .89 |
| Renal symptoms |  |  |  |
| Oliguria (n, %) | 20 (54.1) | 18 (47.4) | 0.56 |
| Anuria (n, %) | 6 (16.2) | 4 (10.5) | 0.52 |
| Hypertension (n, %) | 19 (51.4) | 14 (36.8) | 0.21 |
| Uremic symptoms (n, %) | 18 (48.6) | 15 (39.5) | 0.42 |
| Extra-renal symptoms |  |  |  |
| Skin lesions (n, %) | 3 (8.1) | 5 (13.2) | 0.71 |
| Arthritis (n, %) | 5 (13.5) | 11 (28.9) | 0.1 |
| Hemoptysis (n, %) | 0 | 15 (39.5) | **<0.001** |
| Fever (n, %) | 9 (24.3) | 19 (50) | **0.02** |
| Hemoglobin (g/dL, mean±SD) | 8.8±2.7 | 8.2±2 | 0.19 |
| Total leucocyte (cells*10<sup>9</sup>/L, mean±SD) | 9.3±2.5 | 11.2±4.5 | **0.03** |
| Serum cholesterol (mg/dL, mean±SD, n) | 206.7±61.1 | 155.3±47.9 | **0.003** |
| Serum Albumin (mg/dL, mean±SD) | 3.1±0.5 | 3.1±0.6 | 0.79 |
| 24-hour proteinuria (g/day, median (IQR), n) | 3.7 (2.1-6.7) | 2.1 (0.9-3.5) | **0.001** |
| Serum creatinine (mg/dL, mean±SD) | 7.1±4 | 7.9±4.3 | 0.39 |
| CKD-EPI eGFR (ml/min/1.73m<sup>2</sup>, median (IQR)) | 9 (5.5-16) | 7 (4-10.3) | 0.11 |
| Serum complements (mg/dL) |  |  |  |
| Low C3 (n/N, %) | 12 (32.4) | 6 (16.2) | 0.1 |
| Low C4 (n/N, %) | 1 (2.7) | 0 | 1 |
| Crescents (%) | 74.8± 20.5 | 73± 18.3 | 0.7 |
| Predominant type |  |  |  |
| Cellular (n, %) | 16 (43.2) | 6 (15.8) | **0.009** |
| Fibrocellular or Fibrous (n, %) | 21 (56.8) | 32 (84.2) |  |
| Glomerular lesions |  |  |  |
| Mesangial proliferation Yes (n, %) | 20 (54.1) | 20 (52.6) | 0.9 |
| Neutrophilic infiltration Yes (n, %) | 12 (32.4) | 11 (28.9) | 0.74 |
| Tuft necrosis Yes (n, %) | 5 (13.5) | 14 (36.8) | **0.02** |
| Interstitial |  |  |  |
| IFTA Moderate/Severe Yes (n, %) | 20 (54.1) | 20 (52.6) | 0.9 |
| Vascular |  |  |  |
| Necrosis Yes (n, %) | 1 (2.7) | 2 (5.3) | 0.57 |
| Arterio(lo)sclerosis Yes (n, %) | 26 (70.3) | 21 (55.3) | 0.18 |
| Treatment |  |  |  |
| Immunosuppression Yes (n, %) | 33 (89.2) | 36 (94.7) | 0.38 |
| PLEX Yes (n, %) | 3 (8.1) | 17 (44.7) | **<0.001** |
| Outcomes |  |  |  |
| Serum creatinine at last follow-up (mg/dL, mean±SD, n) | 4±4.2, 18 | 3.1±1.9, 22 | 0.42 |
| eGFRat last follow-up (ml/min/1.73m<sup>2</sup>, median (IQR), n) | 29 (17.8-51.7), 18 | 21.6 (13.2-34.7), 22 | 0.09 |
| ESKD | 20 (54.1) | 17 (44.7) | 0.42 |
| Death | 0 | 5 (13.2) | 0.07 |

Abbreviations: eGFR, estimated glomerular filtration rate (calculated using the CKD-EPI, Chronic Kidney Disease Epidemiology
Collaboration (CKD-EPI) formula); ESKD, end stage kidney disease; C3, complement C3; C4, complement C4; IFTA, interstitial fibrosis and tubular atrophy; PLEX, plasma exchange; ESKD, end stage kidney disease

**Figure 2.**
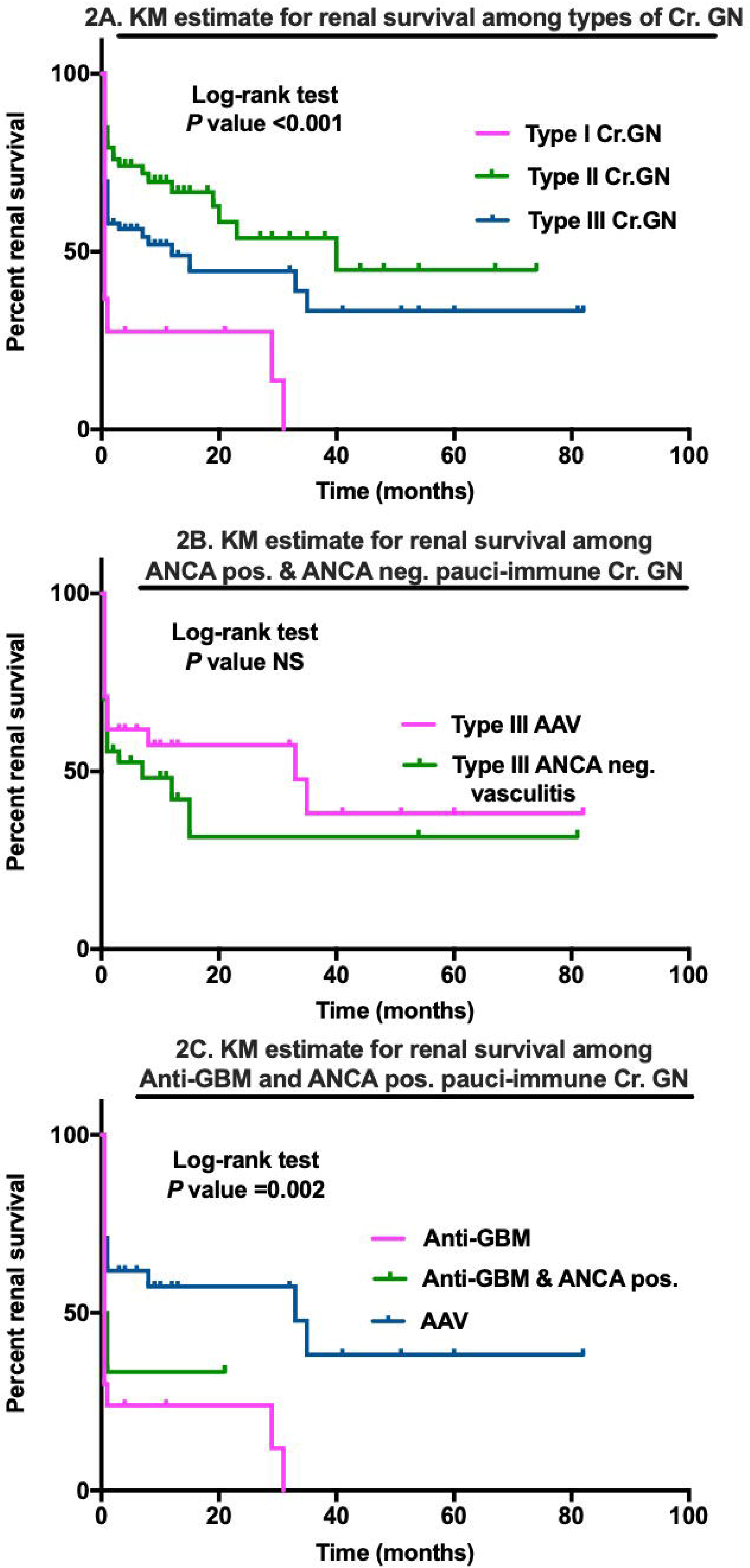
Kaplan-Meier estimates for renal survival in types of crescentic glomerulonephritis & their subgroups

### Double Antibody positive Cr.GN

Among those with type I Cr.GN (n=31), there were 10 ‘double positive’ patients (32.3%) with positive ANCA serologies (anti-MPO 7; anti-PR3 3). Renal survival was significantly worse in this group (p=0.002) compared to AAV but similar to type I Cr.GN (Figure 2).

### Predictors of severe outcomes

We analyzed the risk factors associated with dialysis dependence at discharge from index visit in the entire cohort (Table 6). In an adjusted regression analysis, type I/type III Cr.GN, moderate to severe IFTA, presence of oliguria/anuria and increasing percentage of crescents in renal biopsy were significant predictors of dialysis dependence at discharge. We also analyzed various risk factors which predicted the development of ESKD at follow-up in this cohort. (Table 7). Similarly, in an adjusted regression analysis, type I/type III Cr.GN, moderate to severe IFTA, presence of oliguria/anuria and increasing percentage of crescents in renal biopsy were significant predictors of ESKD at follow-up. Kaplan-Meier estimates of renal survival differed significantly between the three groups (Figure 2) but did not differ in any subgroups (Figure 2). Patient survival was similar in all three groups (Figure 3).

**Table 6.** Significant predictors of dialysis dependence at index visit discharge

| Risk factors | Univariate<br>*P* value | Multivariable regression |  |  |  |
| --- | --- | --- | --- | --- | --- |
|  |  | Exp (B) | 95% C.I. |  | *P* value |
|  |  |  | Lower | Upper |  |
| Gender Males | 0.07 |  |  |  |  |
| Renal symptoms |  |  |  |  |  |
| Oliguria Yes | <0.001 | ] 7.8 | 3.7 | 16.4 | <0.001 |
| Anuria Yes | <0.001 |  |  |  |  |
| Uremic symptoms Yes | <0.001 |  |  |  |  |
| Extra-renal symptoms |  |  |  |  |  |
| Hemoptysis Yes | 0.021 |  |  |  |  |
| Arthritis No | 0.054 |  |  |  |  |
| Hb (g/dL) | 0.009 |  |  |  |  |
| Serum albumin (g/dL) | 0.041 |  |  |  |  |
| Serum cholesterol (mg/dL) | 0.007 |  |  |  |  |
| Serum creatinine (mg/dL) | <0.001 |  |  |  |  |
| CKD-EPI eGFR (ml/min/1.73m<sup>2</sup>) | <0.001 |  |  |  |  |
| Serology |  |  |  |  |  |
| ANA neg. | 0.049 |  |  |  |  |
| Percentage of crescents | <0.001 | 1.03 | 1.01 | 1.05 | 0.003 |
| Glomerular lesions |  |  |  |  |  |
| Mesangial proliferation No | 0.031 |  |  |  |  |
| Vascular |  |  |  |  |  |
| Necrosis Yes | 0.022 | 5.6 | 0.6 | 54 | 0.14 |
| Interstitial |  |  |  |  |  |
| IFTA Moderate/Severe Yes | <0.001 | 2.6 | 1.3 | 5.3 | 0.006 |
| Type I/III Cr. GN | <0.001 | 2.7 | 1.3 | 5.4 | 0.006 |
| Immunosuppression No | 0.002 |  |  |  |  |
| PLEX Yes | 0.007 |  |  |  |  |
| Hemodialysis at index visit discharge Yes | <0.001 |  |  |  |  |

**Table 7.** Significant predictors of ESKD at follow-up

| Risk factors | Univariate<br>*P* value | Multivariable regression |  |  |  |
| --- | --- | --- | --- | --- | --- |
|  |  | Exp (B) | 95% C.I. |  | *P* value |
|  |  |  | Lower | Upper |  |
| Gender | 0.22 |  |  |  |  |
| Renal symptoms |  |  |  |  |  |
| Oliguria Yes | **<0.001** | ] 3.1 | 1.6 | 6.1 | **0.001** |
| Anuria Yes | **0.004** |  |  |  |  |
| Uremic symptoms Yes | **<0.001** |  |  |  |  |
| Extra-renal symptoms |  |  |  |  |  |
| Skin lesions No | **0.038** |  |  |  |  |
| Arthritis No | **0.014** |  |  |  |  |
| Serum creatinine (mg/dL) | **<0.001** |  |  |  |  |
| CKD-EPI eGFR (ml/min/1.73m<sup>2</sup>) | **<0.001** |  |  |  |  |
| Normal C3 | 0.66 |  |  |  |  |
| Normal C4 | **0.04** |  |  |  |  |
| Serology |  |  |  |  |  |
| ANA neg. | **0.014** |  |  |  |  |
| Anti- dsDNA pos. | 0.07 |  |  |  |  |
| ANCA pos. | 0.68 |  |  |  |  |
| Percentage of crescents | **<0.001** | 1.02 | 1.004 | 1.04 | **0.016** |
| Glomerular lesions |  |  |  |  |  |
| Mesangial proliferation No | **0.01** |  |  |  |  |
| Predominant type of crescents |  |  |  |  |  |
| Cellular Yes | 0.28 |  |  |  |  |
| Vascular |  |  |  |  |  |
| Necrosis Yes | **0.022** | 6.2 | 0.6 | 62.4 | 0.12 |
| Interstitial |  |  |  |  |  |
| IFTA Moderate/Severe Yes | **<0.001** | 3.1 | 1.6 | 6.1 | **0.001** |
| Type I/III Cr.GN | **<0.001** | 2.7 | 1.4 | 5.3 | **0.003** |
| Immunosuppression No | **0.003** | 2.1 | 0.7 | 6.3 | 0.18 |
| PLEX Yes | 0.1 |  |  |  |  |
| Hemodialysis at index visit discharge Yes | **<0.001** |  |  |  |  |

**Figure 3.**
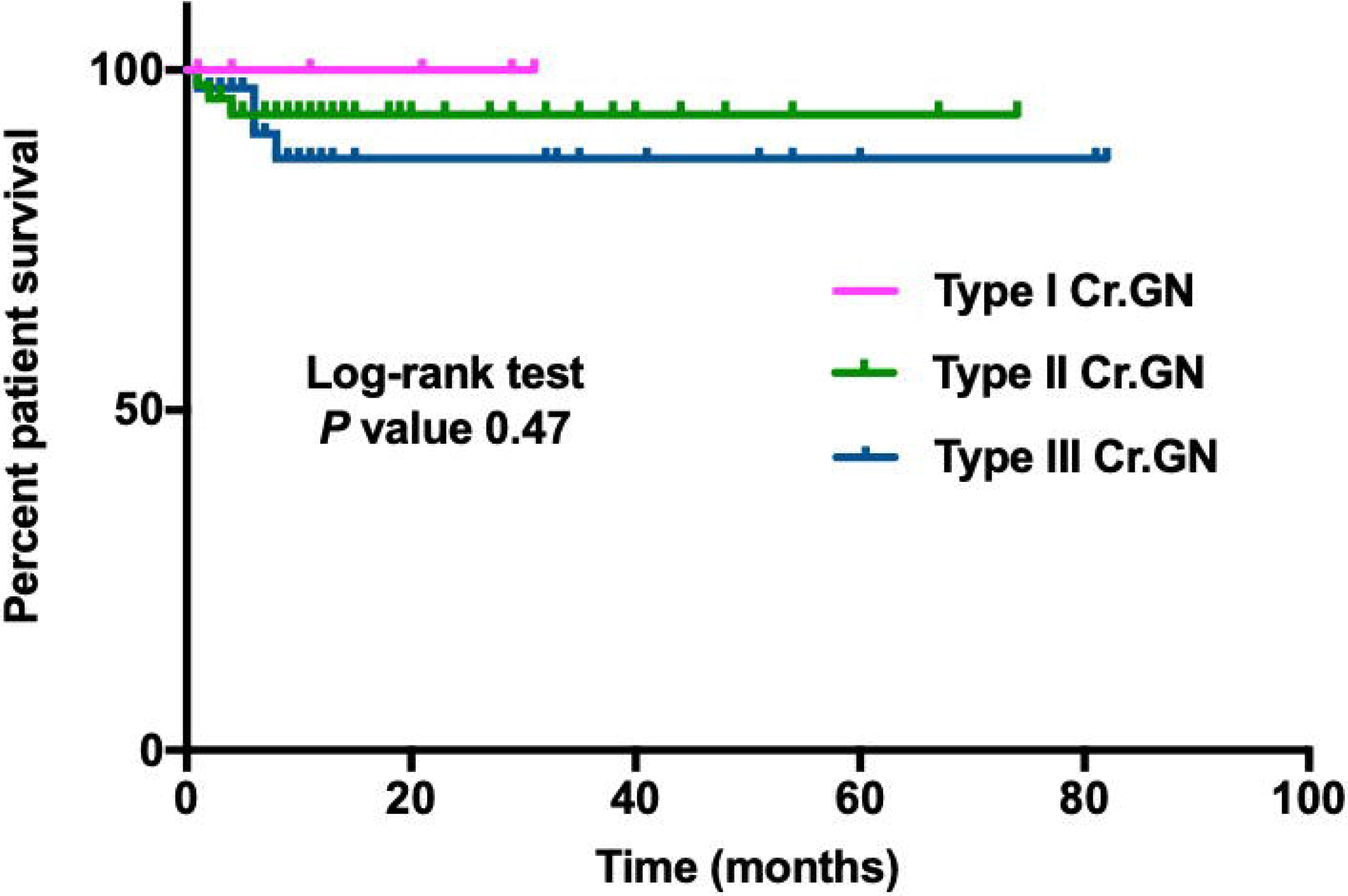
Kaplan-Meier estimate for patient survival in types of crescentic glomerulonephritis

## DISCUSSION

Crescentic GN (Cr.GN) is one of the leading causes of rapidly progressive renal failure. There are a few studies of crescentic GN from South Asia but there is dearth of data on the different types of Cr.GN and their outcomes.

In this study Cr.GN accounted for 2.3% of all biopsies conducted over a period of 10 years, which is comparable to previously reported rates of Cr.GN from India.^11,12^ Type II Cr.GN was found to be the most common type which is unique to Asian continent. Various studies conducted in different parts of the world have shown type III Cr.GN to be the most common.^5,13^ However studies from China, Saudi Arabia and a smaller Indian study have reported type II Cr.GN to be the most common.^2,11,14^ Reasons cited for increased incidence of type II Cr.GN in these parts have been an increased incidence of infections and a higher prevalence of IgA nephropathy. Lupus nephritis (45%) is the most common cause of type II Cr.GN, followed by infection related GN (24%) and IgA nephropathy (23%). This is similar to one of the largest reviews of Cr.GN from China in which lupus nephritis was listed as most common cause of type II Cr.GN (34%) followed by IgA nephropathy (17%).^2^

Gender differences in Cr.GN have mostly been observed in type II Cr.GN with female preponderance. The gender distribution has been variable in type I and type III Cr.GN in different studies. ^2,11,13-15^ As is already known and established in other studies, patients with type I Cr.GN had the most severe renal failure at presentation. However, in this study, more than half of the patients with type II & type III Cr.GN presented with severe renal failure highlighting the severity of presentation in this patient population. These rates are much higher than previously reported.^2,15^

Serum complement C3 levels have been shown to be low in type I and type III Cr.GN in addition to type II Cr.GN. The same was confirmed in our study. However, we found no cases of low C4 levels in type I and III Cr.GN, undersigning the statement that alternate complement pathway has a role in the pathogenesis of these GNs. Our observed rates of ANCA seropositivity in type III Cr.GN are much lower than those reported in other studies.^2,15^An earlier study from India also reports similar rates of ANCA seropositivity.^16^ ANCA negative vasculitis could be attributed to other antibodies such as anti-endothelial cell antibodies, or to cell mediated immune mechanisms which lead to neutrophilic activation.^17^ The high prevalence of ANCA negative vasculitis in this cohort highlights the need for research into its pathogenesis to elucidate factors specific to our population.

Subgroup analysis of type III Cr.GN revealed important differences between AAV and ANCA negative vasculitis groups, being reported for the first time from India. Similar to ours, younger age of onset and lower prevalence of systemic involvement in ANCA negative vasculitis has been reported from other parts of the world.^18-21^ Chen *et al*.^19^ also observed a higher level of proteinuria in this group similar to our study. Although chronic lesions on kidney biopsy were more prevalent in ANCA negative vasculits in these studies, we found a higher prevalence of fibrous/fibrocellular crescents and tuft necrosis in AAV group. Data on renal outcomes has been variable with few studies showing comparable outcomes in the two groups^20,21^ similar to our study and others showing poorer renal outcome in ANCA negative vasculitis.^18,19^

Levy *et al*.^22^ reported a prevalence of ANCA positivity of nearly 30% in type I Cr.GN with predominance of anti-MPO ANCA. They concluded that these patients had a poor prognosis when presenting with severe disease and initially behaved more like anti-GBM disease than vasculitis with low rates of recovery from renal failure. On the other hand SP McAdoo *et al*.^23^ in their retrospective analysis of double antibody positive cases found that such patients shared characteristics of AAV, such as older age distribution and longer symptom duration before diagnosis, and anti-GBM disease, such as severe renal disease and higher frequency of lung haemorrhage at presentation. Despite having more evidence of chronic injury on renal biopsy compared to patients with anti-GBM disease, double-positive patients had a greater tendency to recover from being dialysis dependent after treatment and had intermediate long term renal survival compared to the single-positive patients. Our prevalence of ‘double positive Cr.GN’ was similar to Levi *et al*. with intermediate risk for renal survival.

At the end of follow up period, almost half of the patients had developed ESKD in our study. Renal survival was worst in type I Cr.GN. Similar rates of renal survival were reported by Chen *et al*.^2^ Previously reported rates of renal failure from India varied between 46-60%.^11,16^ Different studies have identified various risk factors for ESKD; sclerosed glomeruli, acute tubular necrosis, vasculopathy,^7^ arteriolar fibrinoid necrosis,^16^ serum creatinine, age, lung involvement, serum CRP,^5^ oliguria, crescents and interstitial inflammation.^2^

## CONCLUSION

We were able to analyse detailed demographic, clinical, serological and pathological features of our cohort. Strengths of our study include its large sample size, the inclusion and comparison of ANCA negative vasculitis as well as ‘double positive Cr.GN’ patients, of which there is no previously reported data from India. We highlight several important clinical practice points - in particular type II Cr.GN may present with a severe renal failure similar to type III Cr.GN, however the response to treatment and outcomes were much more favourable and appropriate treatment should be initiated at earliest. Prevalence of ANCA negative vasculitis was much higher in our population and hence kidney biopsy is mandatory in a case of suspected RPGN with negative serologies. Type I/type III Cr.GN, moderate to severe IFTA, presence of oliguria/anuria and increasing percentage of crescents in renal biopsy were significant predictors of dialysis dependence at index visit or of ESKD at follow-up in our cohort. Our study has also been able to identify areas of further research; search for pathways of alternate complement activation in type I and III Cr.GN and to explore the pathogenesis and factors responsible for increased prevalence of ANCA negative vasculitis in our population. The limitations of the study are that it is a single centre study and it was retrospective in nature.

## Ethics

Approval was obtained from the Institutional Review Board (Silver, Research and Ethics Committee) of the Christian Medical College, Vellore, India (IRB 9090 dated 06.10.2014).

## Data availability

All data underlying the results are available as part of the article and no additional source data are required.

https://www.ebi.ac.uk/biostudies/

## Grant information

This work is supported by the DBT/Wellcome Trust India Alliance Fellowship (grant number IA/CPHE/14/1/501501). S.A. is a DBT/Wellcome Trust India Alliance Early Career Fellow (Clinical and Public Health).

*The funders had no role in study design, data collection and analysis, decision to publish, or preparation of the manuscript*.

The published work was during the Ph.D. of The Tamil Nadu Dr. M.G.R. Medical University.

## Acknowledgements

None

